# Dietary nutrient balances and serum cholesterol: a new approach to an old question

**DOI:** 10.1101/2023.06.14.23291374

**Authors:** Maria Léa Corrêa Leite

## Abstract

The continuing controversy concerning the relationships between diet and serum cholesterol levels highlights the need for innovative analytical approaches to the question. It is now acknowledged that dietary data are compositional in nature, but it is less widely recognised that the same is also true of any variable expressed in concentration units (mg/mL), such as serum cholesterol. Compositional data are parts of a whole and convey essentially relative information, which means they need to be interpreted in terms of the ratios between the individual components of the composition. The various formulations of log-ratio transformations proposed for compositional data analysis provide new variables that can be included in standard regression models as dependent and explanatory variables. Using data from an Italian population-based study, we describe the use of such methods to evaluate the relationships between a two-part composition (non-HDL and HDL cholesterol and their total) and three dietary nutrient compositions, and define multivariate linear regression equations that have one cholesterol log-ratio and the composition total as dependent variables and some macro- and micronutrient log-ratios as explanatory terms. Two alternative models are fitted: one containing the nutrient log-ratios in the form of their simplified expression as orthogonal balances; the other estimating the impact of nutrient pivot balances, which express the relative dominance of each of the parts of the dietary compositions. This approach to investigating the relationship between diet and serum cholesterol allows the simultaneous examination of the effects of non-redundant dietary components on both the quantitative and qualitative aspects of serum cholesterol profiles, and provides insights into some matters concerning public health.

## 1. Introduction

Despite decades of research, there is still no consensus about the relationship between diet and serum cholesterol. The conflicting results of different studies, and the fact that the findings of observational studies are frequently not supported by those of randomised controlled trials, reflect the complexity of dietary data and inadequacies in the way they are analysed. When investigating the role of distinct dietary components, pitfalls such as multicollinearities can lead to unstable and unreliable estimates of effect, which may partially explain the conflicting results of studies based on individual nutrients.

The interdependence of dietary components is intrinsic to their compositional nature but, unfortunately, nutritional researchers have largely ignored the statistical methodologies proposed for the analysis of compositional data. Compositional data are parts of a whole and essentially convey relative information. They are usually recorded as proportions or percentages, although a broader definition has characterised them as vectors of strictly positive measurements in which a variation in one part needs to be interpreted in relation to the other parts, and one of the basic principles of compositional data analysis is that the meaningful information obtainable from the data is contained in the ratios between the individual components of the composition [1].

The variety of log-ratio transformations proposed for compositional data analysis provide new variables that can be included in standard regression models, and we have previously described how to apply log-ratio transformations to dietary compositions using their particular expression as *balances* [2,3]. We have also discussed the interpretative implications of using different log-ratios as explanatory variables in regression models, and shown that applying them to dietary data has numerical and conceptual advantages over traditional analytical methods [4].

The aim of this note is to describe the application of log-ratio methodologies when evaluating the relationships between dietary macro- and micronutrient compositions and serum cholesterol profiles, and discuss some of the insights that this may provide. As concentration units (mg/mL) indicate compositional information (parts of a whole), we also describe regression models in which both the independent variables (dietary data) and the dependent variables (serum cholesterol profiles) are compositional.

## 2. Methods

### 2.1. Log-ratio transformations for compositional data

Various formulations of log-ratios have been proposed as a means of analysing compositional data and one of these is the isometric log-ratio (ilr) transformation [5]. Given a D-part composition (x_1_,x_2_,…,x_D_), the ilr-transformation consists of orthogonally decomposing the parts into non-overlapping subgroups and representing their relationships. One way of defining ilr(s) in their particular form of *balances* is to use sequential binary partition as a bifurcating tree: the parts of a composition are successively and hierarchically split into two groups until all of the groups have a single part. At each of the D-1 steps required to complete the partition, a generic balance is defined as the orthonormal log-ratio of the geometric mean of each group of parts:

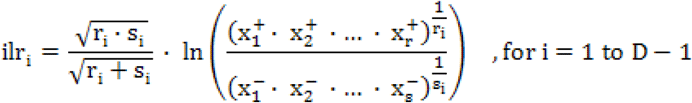

in which the square root coefficient represents the normalising constant, and r and s are respectively the number of parts in the numerator (x^+^) and the number of parts in the denominator (x^-^).

An alternative choice of partition has been proposed [6] as

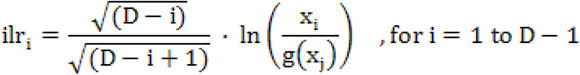

in which g(x_j_) is the geometric mean of the compositional parts of j=i+1 to D. The particular characteristic of this partition is that the first new variable (ilr_1_) captures all of the relevant information concerning compositional part x_1_, and is thus called the *pivot balance* [7]. This special balance expresses the *relative dominance* of the part in the numerator over the other compositional parts. A number of D sets of D-1 ilr(s) are defined, each of which has a different compositional part in the numerator of the pivot balance.

It has also been proposed [8] that removing the normalising constant (and consequently reducing the orthonormality of the log-contrasts to simple orthogonality) would enhance the interpretability of the results of regression analyses involving ilr-balances.

For the sake of simplicity, let us consider the example of a four-part composition (x_1_,x_2_,x_3_,x_4_) for which a given choice of partition produces the following set of three orthogonal balances:

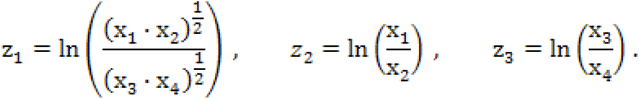

In a regression equation that simultaneously includes z_1_,z_2_ and z_3_ as explanatory variables, the regression coefficient relating to z_1_ represents the estimated variation in the response associated to increases in z_1_ while keeping z_2_ and z_3_ constant. This coefficient therefore represents the expected change in outcome when z_1_ increases in such a way that x_1_ and x_2_ increase by a common factor and x_3_ and x_4_ decrease by a common factor.

Alternatively, the set of balances of which the first is the simplified (orthogonal rather than orthonormal) x_1_-pivot balance is:

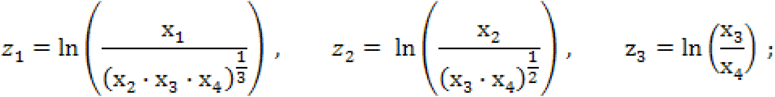

and that defining the simplified x_2_-pivot balance is

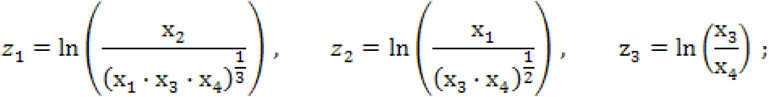

and so on.

Note that z_1_ captures all of the information relating to the component in the numerator that does not appear in the other balances. When including z_1_, z_2_ and z_3_ as explanatory variables in regression equations, at each time, the z_1_-coefficients represent the expected change in response related to increases in the pivot balance made in such a way that the part in the numerator increases against all of the other components, which decrease by a common factor.

As the pivot approach requires a number of runs that is equal to the number of components in the composition, it can involve a considerable amount of work in the case of compositional vectors with a large number of parts. One way of overcoming this drawback is to use the additive log-ratio (alr) transformation, which involves dividing each D-1 component by one that is arbitrarily chosen (for example the last). Using our example of a four-part composition:

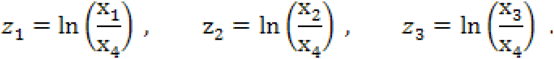

It has been shown that alr and simplified (orthogonal rather than orthonormal) pivot balances are explanatory equivalents [9], and that using alr transformations as predictors and only two runs leads to the same result as the pivot approach [4].

### 2.2. Serum cholesterol profiles and dietary balances

The results shown below consider a two-part serum cholesterol composition (high-density lipoprotein [HDL] and non-HDL cholesterol) and their sum (total cholesterol [TC]), and fit two multivariate linear regression models with ln(TC) and the log-ratio ln(non-HDL/HDL) as dependent variables after defining the nutrient balances of three dietary compositions: an eight-part macronutrient, a nine-part vitamin, and a six-part mineral composition. For illustrative purposes, the first model includes the orthogonal balances of each dietary composition as explanatory variables, and the second includes their pivot balances as explanatory variables. In addition to dietary variables (total energy and fibre intake, and the macro- and micronutrient balances), the models include terms for gender, age, smoking, alcohol consumption, practising sport and television watching time.

The approach is illustrated using data coming from the Italian Bollate Eye Study[10], a population-based study of subjects aged 40-74 years that was carried out in 1992-1993, in which dietary habits were assessed by means of a food-frequency questionnaire, and mean daily nutrient intakes were calculated using the food compositional database compiled for epidemiological studies in Italy [11]. The study was approved by the Ethics Committee of Italy’s National Research Council (CNR).

## 3. Results

Table 1 shows the sign matrix on which the partition applied to the eight-part macronutrient composition was based. Accordingly, the second balance is the (simplified) balance between proteins and carbohydrates in kcal/day, calculated as

**Table 1.** Sequential binary partitions of an eight-part macronutrient composition. At each step, +1 indicates that the part is assigned to the first group (numerator), -1 indicates that it is assigned to the second group (denominator), and 0 indicates that it is not involved in the partition at this step.

| Step | Macronutrients |  |  |  |  |  |  |  | r | s |
| --- | --- | --- | --- | --- | --- | --- | --- | --- | --- | --- |
|  | AP | VP | ST | SS | SFA | MUFA | n-6LA | n-3ALA |  |  |
| 1 | -1 | -1 | -1 | -1 | +1 | +1 | +1 | +1 | 4 | 4 |
| 2 | +1 | +1 | -1 | -1 | 0 | 0 | 0 | 0 | 2 | 2 |
| 3 | +1 | -1 | 0 | 0 | 0 | 0 | 0 | 0 | 1 | 1 |
| 4 | 0 | 0 | +1 | -1 | 0 | 0 | 0 | 0 | 1 | 1 |
| 5 | 0 | 0 | 0 | 0 | +1 | -1 | -1 | -1 | 1 | 3 |
| 6 | 0 | 0 | 0 | 0 | 0 | +1 | -1 | -1 | 1 | 2 |
| 7 | 0 | 0 | 0 | 0 | 0 | 0 | +1 | -1 | 1 | 1 |

AP: animal proteins; VP: vegetable proteins; ST: starches; SS: simple sugars; SFA: saturated fatty acids; MUFA: monounsaturated fatty acids; LA: linoleic acids; ALA: alpha-linolenic acids.

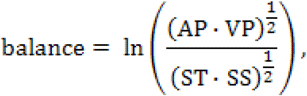

where AP is animal proteins, VP vegetable proteins, ST starches, and SS simple sugars.

The regression analyses of the serum cholesterol variables against the dietary and control variables are based on data relating to 1090 non-diabetic subjects (547 men and 543 women). Tables 2 and 3 show the dietary component results obtained using the alternative approaches of nutrient orthogonal balances (Tab. 2) and nutrient pivot balances (Tab. 3). The hierarchical manner in which the orthogonal balances were defined can easily be deduced by examining the way in which they were designed (Tab. 2, first column). The use of multivariate models makes it possible to obtain overall tests (the F-tests in the last column of the two tables) that denote the impact of each nutrient balance on the serum cholesterol composition as a whole, but how are the B-coefficients interpreted?

**Table 2.** Multivariate linear regression coefficients (B) and standard errors (SE) of serum cholesterol profiles (total cholesterol and non-HDL/HDL balance) in relation to mean daily energy and fibre intake and dietary macro- and micronutrient balances. Model 1: Orthogonal balances.

| Dietary variables: | Ln (total cholesterol) |  | Ln (non-HDL/HDL) |  | F-test<sub>(2,1049)</sub> |
| --- | --- | --- | --- | --- | --- |
|  | B (SE) | P_value | B (SE) | P_value | P_value |
| Total energy [ln(kcal)] | -0.108 (0.046) | 0.018 | 0.061 (0.097) | 0.530 | 0.005 |
| Fibre [ln(mg)] | 0.091 (0.043) | 0.037 | -0.026 (0.092) | 0.779 | 0.028 |
| *Macronutrients* |  |  |  |  |  |
| Fats vs proteins and carbohydrates | 0.021 (0.036) | 0.560 | 0.042 (0.077) | 0.586 | 0.813 |
| Proteins vs carbohydrates | -0.115 (0.053) | 0.031 | -0.046 (0.113) | 0.683 | 0.063 |
| Animal vs vegetable proteins | 0.045 (0.037) | 0.223 | -0.041 (0.079) | 0.606 | 0.177 |
| Starches vs simple sugars | -0.001 (0.023) | 0.976 | 0.020 (0.049) | 0.687 | 0.882 |
| Saturated vs unsaturated fats | -0.083 (0.030) | 0.006 | -0.134 (0.064) | 0.036 | 0.018 |
| Monounsaturated vs polyunsaturated fats | 0.033 (0.024) | 0.159 | 0.023 (0.050) | 0.648 | 0.345 |
| n-6 linoleic vs n-3 alpha-linolenic acids | -0.001 (0.026) | 0.979 | -0.073 (0.056) | 0.187 | 0.282 |
| *Vitamins* |  |  |  |  |  |
| Vitamin B vs vitamin D, E, C, beta-carotene | 0.039 (0.044) | 0.377 | 0.009 (0.092) | 0.925 | 0.608 |
| B<sub>1</sub>, B<sub>2</sub>, B<sub>3</sub> vs B<sub>6</sub>, B<sub>9</sub> (folates) | -0.006 (0.067) | 0.925 | 0.284 (0.143) | 0.047 | 0.051 |
| B<sub>3</sub> vs B<sub>1</sub>, B<sub>2</sub> | 0.046 (0.035) | 0.180 | 0.185 (0.073) | 0.012 | 0.041 |
| B<sub>1</sub> vs B<sub>2</sub> | -0.041 (0.044) | 0.348 | -0.103 (0.092) | 0.265 | 0.497 |
| B<sub>6</sub> vs B<sub>9</sub> (folates) | -0.062 (0.038) | 0.100 | -0.174 (0.080) | 0.030 | 0.081 |
| Vitamin D vs vitamin E, C, beta-carotene | 0.025 (0.015) | 0.105 | 0.016 (0.033) | 0.615 | 0.242 |
| Vitamin C vs vitamin E, beta-carotene | 0.001 (0.020) | 0.965 | 0.047 (0.043) | 0.271 | 0.438 |
| Vitamin E vs beta-carotene | -0.025 (0.023) | 0.291 | -0.073 (0.050) | 0.139 | 0.320 |
| *Minerals* |  |  |  |  |  |
| Na, K vs Ca, P, Fe, Z | -0.124 (0.065) | 0.056 | 0.060 (0.138) | 0.665 | 0.034 |
| Ca, P vs Fe, Z | -0.014 (0.058) | 0.807 | -0.130 (0.124) | 0.293 | 0.532 |
| Na vs K | 0.074 (0.032) | 0.022 | -0.019 (0.068) | 0.786 | 0.014 |
| Ca vs P | -0.004 (0.048) | 0.942 | 0.227 (0.103) | 0.027 | 0.027 |
| Fe vs Z | -0.009 (0.041) | 0.825 | -0.182 (0.088) | 0.039 | 0.065 |

Na: sodium; K: potassium; Ca: calcium; P: phosphorus; Fe: iron; Z: zinc.
The model also included terms for gender, age, practising sport, television watching time, smoking, and alcohol consumption.

**Table 3.** Multivariate linear regression coefficients (B) and standard errors (SE) of serum cholesterol profiles (total cholesterol and non-HDL/HDL balance) in relation to mean daily energy and fibre intake and dietary macro- and micronutrient balances. Model 2: Pivot balances (relative dominances).

| Dietary variables: | Ln (total cholesterol) |  | Ln (non-HDL/HDL) |  | F-test (2,1049) |
| --- | --- | --- | --- | --- | --- |
|  | B (SE) | P_value | B (SE) | P_value | P_value |
| Total energy [ln(kcal)] | -0.108 (0.046) | 0.018 | 0.061 (0.097) | 0.530 | 0.005 |
| Fibre [ln(mg)] | 0.091 (0.043) | 0.037 | -0.026 (0.092) | 0.779 | 0.028 |
| *Macronutrients* |  |  |  |  |  |
| Animal proteins vs other macronutrients | -0.018 (0.036) | 0.621 | -0.074 (0.076) | 0.328 | 0.619 |
| Vegetable proteins vs other macronutrients | -0.108 (0.060) | 0.070 | 0.007 (0.126) | 0.956 | 0.089 |
| Starches vs other macronutrients | 0.052 (0.041) | 0.208 | 0.032 (0.087) | 0.709 | 0.422 |
| Simple sugars vs other macronutrients | 0.053 (0.022) | 0.018 | -0.007 (0.047) | 0.877 | 0.014 |
| Saturated fats vs other macronutrients | -0.078 (0.031) | 0.013 | -0.124 (0.066) | 0.063 | 0.038 |
| Monounsaturated fats vs other macronutrients | 0.066 (0.029) | 0.024 | 0.078 (0.062) | 0.210 | 0.078 |
| n-6 Linoleic acid vs other macronutrients | 0.017 (0.031) | 0.587 | -0.030 (0.066) | 0.654 | 0.581 |
| n-3 Alpha-linolenic acid vs other macronutrients | 0.016 (0.031) | 0.610 | 0.117 (0.065) | 0.072 | 0.168 |
| *Vitamins* |  |  |  |  |  |
| Thiamin vs other vitamins | -0.058 (0.050) | 0.244 | -0.099 (0.106) | 0.351 | 0.476 |
| Riboflavin vs other vitamins | 0.023 (0.050) | 0.642 | 0.107 (0.106) | 0.315 | 0.601 |
| Niacin vs other vitamins | 0.052 (0.046) | 0.257 | 0.281 (0.097) | 0.004 | 0.013 |
| Vitamin B6 vs other vitamins | -0.051 (0.059) | 0.388 | -0.314 (0.126) | 0.013 | 0.037 |
| Folates vs other vitamins | 0.073 (0.044) | 0.098 | 0.034 (0.093) | 0.718 | 0.206 |
| Vitamin D vs other vitamins | 0.015 (0.013) | 0.220 | 0.014 (0.027) | 0.591 | 0.465 |
| Vitamin E vs other vitamins | -0.043 (0.039) | 0.264 | -0.105 (0.082) | 0.201 | 0.389 |
| Beta-carotene vs other vitamins | 0.006 (0.018) | 0.732 | 0.042 (0.039) | 0.279 | 0.532 |
| Vitamin C vs other vitamins | -0.017 (0.023) | 0.457 | 0.039 (0.049) | 0.414 | 0.261 |
| *Minerals* |  |  |  |  |  |
| Sodium vs other minerals | 0.012 (0.015) | 0.414 | 0.012 (0.031) | 0.708 | 0.713 |
| Potassium vs other minerals | -0.136 (0.063) | 0.031 | 0.048 (0.134) | 0.717 | 0.018 |
| Calcium vs other minerals | 0.020 (0.033) | 0.533 | 0.149 (0.070) | 0.033 | 0.088 |
| Phosphorus vs other minerals | 0.027 (0.074) | 0.711 | -0.307 (0.158) | 0.052 | 0.034 |
| Iron vs other minerals | 0.029 (0.046) | 0.529 | -0.142 (0.099) | 0.150 | 0.109 |
| Zinc vs other minerals | 0.047 (0.062) | 0.446 | 0.228 (0.132) | 0.084 | 0.208 |

The model also included terms for gender, age, practising sport, television watching time, smoking, and alcohol consumption.

As in the case of any multiple regression framework, one B-coefficient represents the change in response associated with a one unit change in the corresponding nutrient balance, while keeping all of the other terms in the equation constant. Addition on the log scale is equivalent to multiplication on the natural scale and, given that both the dependent and the independent variables are logged and that B = ln(*e*^*B*^) and 1 = ln(*e*), the expected multiplicative change in the response (TC or the non-HDL/HDL ratio) is given by an *e*^*B*^ factor when the relative nutrient ratio is multiplied by *e* (approximately 2.7). It is worth noting that using binary logarithms (base 2) of the nutrient ratios is often useful as it corresponds to the effect of doubling the ratio. Interpretation can also be improved by examining the signs of the two B-coefficients of each explanatory nutrient balance: the coefficient related to the quantitative aspect (TC) of the serum cholesterol profile and the coefficient related to its qualitative aspect (the non-HDL/HDL balance). Generally, when the two coefficients have the same sign, the variations in the cholesterol profile should be mainly attributed to the non-HDL component but, when they have different signs, it should be mainly attributed to the HDL component. Note, for example, that the increases in the dietary *saturated vs unsaturated fats* balance inversely related to the serum non-HDL/HDL balance, the decreases in which were probably due to the reduction in the non-HDL component as the TC-related coefficient was also negative.

Table 3 shows the results of the model including the alr(s) - defined for each of the three dietary compositions - as explanatory variables, and so they can be interpreted as pivot balances. A second run, in which the component in the denominator of the alr(s) was changed, made it possible to obtain the estimates relating to the compositional part that was in the denominator of the alr(s) in the first run. In this context, the regression coefficients express the impact of increasing each nutritional component against reductions in the other compositional parts by a common factor (relative dominance) in such a way that the ratio is multiplied by an *e*-factor. As the different formulations of the log-ratios represent reparametrisations of the same model, the estimates related to the covariates common to the two models and the measures of their performance were precisely the same. Note, for example, that the estimates relating to total energy and fibre intake are exactly the same in the two models.

## 4. Discussion

This paper describes a new approach to investigating the relationship between diet and serum cholesterol profiles that uses all of the available dietary information and thus allows a comprehensive overview and ensures better control of confounding. The determination of balances that are orthogonal (or linearly independent) provides a means of defining new variables that convey non-redundant information. The transformed data can be analysed by means of any of the most popular statistical software.

The isocaloric effects of macronutrients are usually evaluated by means of substitution analysis, a statistical technique based on differences between regression coefficients that tries to mimic studies comparing diets that have different macronutrient compositions. For example, it is possible to estimate the effect of replacing a certain amount of calories coming from saturated fatty acids (SFA) with those coming from polyunsaturated fatty acids (PUFA) or monounsaturated fatty acids (MUFA). However, in the real world, it is impossible to eat SFA, MUFA or PUFA alone as they are generally all present in all fatty foods (what varies is their proportions): for example, olive oil is the principal source of MUFA but in our data it is also the third most important source of SFA after dairy products and meat, and so enriching a diet with olive oil leads to a change in the proportions of SFA and MUFA rather than the replacement of SFAs by MUFAs. Measuring the effects of nutrients in terms of the variations in their proportional ratios therefore seems to reflect real mechanisms more closely.

Furthermore, substitution analyses of types of fats that are based on the replacement of their food sources may fail to consider the inevitable changes in micronutrient contents [12], whereas our approach simultaneously examines the proportional variations in both macro- and micronutrients and is a better reflection of the real-life complexity of dietary changes.

The choice of non-HDL as a component of the serum cholesterol profile is supported by evidence showing that its performance in predicting cardiovascular risk is better than that of low-density lipoprotein (LDL) cholesterol [13]. Non-HDL cholesterol refers to all Apo-B lipoproteins, including LDL, very low-density lipoprotein (VLDL), intermediate-density lipoprotein (IDL), lipoprotein(a), and chylomicron remnants [14] and, as “everything that is not HDL is atherogenic”, it has been suggested that the term non-HDL could be replaced by *atherogenic cholesterol* [15].

Although limited by the cross-sectional nature of the data and bearing in mind that the main aim of this note is to describe an analytical approach, our findings suggest some insights that are interesting from a public health perspective and merit a brief discussion. We found that the increase in the dietary saturated/unsaturated fats ratio is inversely related to the most atherogenic lipid fractions, an apparently surprising finding that conflicts with those of studies showing that an absolute increase in SFA intake is positively associated with serum LDL-C levels, which can be reduced by replacing SFA with PUFA [16,17]. However, other studies based on dietary fatty acid ratios have shown that increasing the PUFA/SFA ratio leads to a worse serum lipid profile [18,19]. Taken together with evidence from prospective observational studies and randomised controlled trials that have failed to confirm the harmful effect of dietary SFA and the beneficial effect of dietary unsaturated fatty acids [20-22], our finding strengthens growing opposition to the conventional dietary recommendation of low SFA and high unsaturated fats consumption.

Our findings also indicate that dietary calcium (Ca)/phosphorus (P) (im)balance is positively related to the non-HDL/HDL ratio or, more precisely, that a worse cholesterol profile is directly related to the intake of Ca and inversely related to the intake of P. This may help to explain the apparent paradox arising from the findings of studies showing that, although dairy Ca may have a beneficial effect [23,24], supplementary Ca may adversely affect serum lipid profiles [25,26]. Our data concerning the role of the Ca/P balance suggest that this difference may be at least partially due to the disrupted balance caused by supplementary Ca insofar as dairy products are rich in both Ca and P.

In conclusion, this paper describes an innovative method of handling explanatory dietary data by flexibly decomposing all of the available dietary information into non-overlapping components whose effects on health outcomes, which may well have a compositional nature, can be easily examined and interpreted.

## Data Availability

All data produced in the present study are available upon reasonable request to the authors

